# Identifying patients with rapid progression from hormone-sensitive to castration-resistant prostate cancer: a retrospective study

**DOI:** 10.1101/2022.10.23.22281406

**Authors:** Chenxi Pan, Yi He, He Wang, Yang Yu, Lu Li, Lingling Huang, Mengge Lyu, Weigang Ge, Bo Yang, Yaoting Sun, Tiannan Guo, Zhiyu Liu

**Affiliations:** Department of Urology, The Second Hospital of Dalian Medical University, No.467 Zhongshan Road, Dalian, Liaoning, China; Key Laboratory of Structural Biology of Zhejiang Province, School of Life Sciences, Westlake University, No.18 Shilongshan Road, Hangzhou, Zhejiang Province, China; School of Life Sciences, Westlake University, No.18 Shilongshan Road, Hangzhou, Zhejiang Province, China; Institute of Basic Medical Sciences, Westlake Institute for Advanced Study, No.18 Shilongshan Road, Hangzhou, Zhejiang Province, China; Westlake Omics (Hangzhou) Biotechnology Co., Ltd., Hangzhou, Zhejiang, China

**Keywords:** hormone-sensitive prostate cancer, PCT-PulseDIA, proteomics, risk stratification, castration-resistance, machine learning

## Abstract

**Background:** Prostate cancer (PCa) is the second most prevalent malignancy and the fifth cause of cancer-related deaths in men. A crucial challenge is identifying the population at risk of rapid progression from hormone-sensitive PCa (HSPC) to the lethal castration-resistant PCa (CRPC).

**Methods:** We collected 78 HSPC biopsies and measured their proteomes using pressure cycling technology and a pulsed data-independent acquisition pipeline. The proteomics data and clinical metadata were used to generate models for classifying HSPC patients and predicting the development of each case.

**Results:** We quantified 7,961 proteins using the HSPC biopsies. A total of 306 proteins were differentially expressed between patients with a long- or short-term progression to CRPC. Using a random forest model, we identified ten proteins that significantly discriminated long-from short-term cases, which were used to classify PCa patients with an 86% accuracy. Next, two clinical parameters (Gleason sum and total PSA) and five proteins (DPT, ARGEF1, UTP23, CMAS, and ANAPC4) were found to be significantly associated with rapid disease progression. A nomogram model using these seven features was generated for stratifying patients into groups with significant progression disparities (*p*-value = 5.2 × 10^−9^).

**Conclusion:** We identified proteins associated with a fast progression to CRPC and an unfavorable prognosis. Based on these proteins, our machine learning and nomogram models stratified HSPC into high- and low-risk groups and predict their prognoses. These tools may aid clinicians in predicting the progression of patients, guiding individualized clinical management and decisions.

## Introduction

Prostate cancer (PCa) is the second most prevalent malignancy in males and the fifth leading cause of cancer-related death globally^1^. With the implementation of prostate-specific antigen screening (PSA) and the aggravation of population aging, after 2012, the PCa incidence and cancer-related mortality in China began to climb^2^. Regarding the risk categories of PCa, locally advanced PCa and metastatic PCa have significantly higher 10- and 15-year mortality rates than other categories^3^. Androgen deprivation therapy (ADT) combined with androgen blocking is frequently beneficial to patients with locally advanced and metastatic PCa during the initial treatment stage^4^. However, almost all hormone-sensitive prostate cancers (HSPC) progress to castration-resistant prostate cancers (CRPC) within five years, with only 5-10% of patients remaining alive ten years after initiating ADT^5^.

Due to its heterogeneity, PCa has a complex disease spectrum, ranging from clinically indolent subtypes to aggressive ones. The progress span to CRPC varies significantly among patients; however, limited research has been conducted to explore this. Multiple randomized controlled phase-III trials, including CHAARTED^6^ and LATITUDE^7^, demonstrated that when HSPC patients are found to have either a long- or short-term progression to CRPC before initiating treatment, it is possible to implement an early and appropriate follow-up strategy, thereby optimizing treatment regimens. Therefore, it is urgent to predict and identify HSPC patients with a long- or short-term progression to CRPC.

The ability to understand the genetics behind large next-generation sequencing datasets has greatly improved. However, not all the genetic or transcriptomic aberrations of PCa are translated into the proteome. Specifically, Latonen et al. reported that gene copy numbers, DNA methylations, and RNA expression levels do not reliably predict the proteomics changes of PCa, especially CRPC^8^. By quantifying and validating large numbers of proteins, these findings indicate that proteomics could be a more promising approach for identifying the molecular mechanisms underlying HSPC progression to CRPC.

In recent years, pressure cycling technology (PCT) and pulsed data-independent acquisition (PulseDIA) have enabled the in-depth and fast identification of proteins in little amounts within tissues, not only fresh frozen^9^ but also formalin-fixed and paraffin-embedded (FFPE) ones^10^. This technology is particularly suitable for measuring the proteomes of trace amounts of clinical biopsies. Through the use of FFPE clinical biopsies tissue, this technology tackles the problem that the data collecting period is too long owing to the collection of clinical prognosis information, therefore considerably speeding up the understanding of major diseases or crucial phases of clinical research.

This study explored the proteomics differences between HSPC patients with a long- or short-term progression to CRPC. To this aim, we used the PCT-PulseDIA pipeline to analyze the proteomes of pre-ADT PCa biopsies. In addition, the proteomes of HSPC with significantly distinct progression were retrospectively evaluated using clinical data. Finally, we generated a machine learning model for classifying patients as undergoing a long- or short-term progression to CRPC and a nomogram model for predicting the advancement of HSPC.

## Materials and methods

### Patient recruitment and sample collection

A total of 78 HSPC patients with complete follow-up information were recruited between January 2014 and July 2021 in the Second Hospital of Dalian Medical University (median [min, max] follow-up time (months): 9.00 [2.00, 65.00]). The Clinical characteristic are given in sTable1. This research was approved by the ethical committee of the Second Hospital of Dalian Medical University with the Declaration of Helsinki. The study was registered in the Chinese Clinical Trial Register (ChiCTR2100054836), and all patients signed a written informed consent before participation.

Locally advanced PCa samples were classified according to the European Association of Urology Prostate Cancer Guideline as any PSA, cT3-4 or cN+, any International Society of Urological Pathology (ISUP) grade or Gleason sum (GS), while metastatic PCa was defined as cM1 disease based on CT and bone scans^11,12^. Accordingly, the patients from our study population were diagnosed with locally advanced (N = 4) or metastatic disease (N = 74) based on histopathological biopsies. Subsequently, all patients were treated with ADT (luteinizing hormone-releasing hormone agonists: goserelin 3.6 mg, once every 28 days, one dose each time, or leuprorelin 3.75 mg, once every 28 days, one dose each time, subcutaneous injection in numerous areas of the upper arm, belly, and buttocks) and with anti-androgen (bicalutamide 50 mg s.i.d. or flutamide 250 mg t.i.d. taken orally). This was the initial and only therapy before progression. Those patients that progressed to CPRC met the following criteria: 1) serum testosterone level < 50 ng/dl, or 1.7 nmol/L; 2) PSA progression: PSA value > 2.0 ng/mL, interval 1 week, three times higher than the baseline level > 50 percent. All patients were followed up till their advancement to CRPC. Patients with cardiovascular diseases, autoimmune diseases, other malignancies, or deceased due to other causes were excluded. The GS was used to evaluate each PCa case’s malignancy and annotate primary and secondary patterns.

### Proteomics sample preparation and data analysis

The pre-ADT biopsies were punched (diameter 1 mm) from the FFPE blocks at the histopathological sites, and the pathologists evaluated the primary Gleason patterns. A total of 78 tissue core samples were collected and then assigned to two groups: a discovery set (n=16) and a modeling set (n=62). These samples were then analyzed using the PCT-PulseDIA technology.

The PCT-assisted proteomics sample preparation procedures followed our previously published workflows^13^. In brief, about 0.2 mg of FFPE punches were dewaxed with heptane, hydrated with ethanol, and then underwent acidic hydrolysis by 0.1% formic acid (FA, Thermo Fisher Scientific, USA) and basic hydrolysis by 0.1 M Tris-HCl (pH = 10.0). Samples were next lysed using a 6 M urea/2 M thiourea buffer (Sigma, USA), reduced by tris (2 carboxyethyl) phosphine (Sigma, USA), and alkylated by iodoacetamide (Sigma, USA). The lysates were then digested using PCT by a mix of Lys-C and trypsin (Hualishi Tech. Ltd., China). Finally, the PCT-assisted digestion reaction was stopped by trifluoroacetic acid and cleaned by C18.

A total of 400 ng peptides were injected and separated along a 45 min liquid chromatography gradient (from 3 to 28% buffer B – see below for its composition) at a flow rate of 300 nL/min (precolumn: 3 μm, 100 Å, 20 mm × 75 μm i.d.; analytical column: 1.9 μm, 120 Å, 150 mm × 75 μm i.d.). Buffer A was mass spectrometry-grade water containing 2% acetonitrile and 0.1% FA; buffer B was acetonitrile containing 2% H_2_O and 0.1% FA. The peptides were then analyzed by a Q Exactive HF hybrid Quadrupole-Orbitrap (Thermo Fisher Scientific, USA) using the PulseDIA mode with four pulses, as previously described^14^.

To analyze the PulseDIA data, we generated an experimental spectral library for the PCa tissues. We combined the cleaned peptides from the discovery dataset into a mixture containing 100 µg peptides. The peptide pool was then separated using Thermo Ultimate Dinex 3000 (Thermo Fisher Scientific, USA) with an XBridge Peptide BEH C18 column (300 Å, 5 µm x 4.6 mm x 250 mm) (Waters, Milford, MA, USA) and a 60 min gradient. Finally, we collected 20 peptide fractions. The fraction data were acquired using data-dependent acquisition (DDA). Four fractions were randomly selected and analyzed by MS twice. Spectronaut™ Pulsar X (version 14.6, Biognosys, Switzerland) was used to generate the spectral library^15^. The DDA files were searched by Pulsar against a human Swiss-Prot FASTA database (downloaded on 2020-01-22), including 20,367 protein sequences; the settings were left to their default values. The established library comprised 143,347 peptide precursors, 115,257 modified peptides, and 9,644 proteins. Next, PulseDIA files were analyzed using Spectronaut with default settings.

### Machine learning model generation and evaluation

To generate a model for stratifying the patients with long- or short-term progression, we screened the differentially expressed proteins (DEPs) from the discovery set having *p*-value < 0.01 and fold-change > 2. The discovery set included long- (L, n=8) and short-term (S, n=8) progression cases.

To generate a machine learning model and a nomogram, the resulting 306 DEPs were preserved in the modeling set, and their missing values were imputed by zero. The modeling set was separated into a training set of 41 samples (∼2/3), and an independent testing set of 21 samples (∼1/3). According to the median progression time (nine months) from HSPC to CRPC, the label of samples was further divided into an S group and an L group.

We conducted an analysis of variance (ANOVA) on the training set for each DEP, selected the significant proteins whose corresponding *p*-values were less than 0.05 according to the ANOVA, constructed a random forest (RF) model using the ten significant proteins based on the training set, and validated the RF model using the independent testing set. Specifically, we created 500 trees, set the node size hyperparameter to one, selected the Gini index as the importance metric for the variables, and left all the other hyperparameters at their default values. We used “mlr3” R package for the above machine learning analysis.

### Establishing a nomogram for predicting the disease progression

The multivariable Cox regression (R package “survival”) was used to determine the prognostic significance of 16 features, comprising six clinical characteristics (age, T-stage, N- stage, M-stage, total PSA (tPSA), GS) and ten proteins from the RF model (Protein name (gene name): Rho guanine nucleotide exchange factor 1 (ARHGEF1), Protein Hook homolog 3 (HOOK3), ATP-dependent RNA helicase DDX24 (DDX24), Dermatopontin (DPT), N- acylneuraminate cytidylyltransferase (CMAS), Glucose 1,6-bisphosphate synthase (PGM2L1), rRNA-processing protein UTP23 homolog (UTP23), Protein FRA10AC1 (FRA10AC1), Anaphase-promoting complex subunit 4 (ANAPC4), Collagen alpha-3(V) chain (COL5A3)). In addition, the R package “forestplot” was used to visualize each variable (*p*-value, hazard ratio (HR), and 95% confidence interval (CI)).

Next, the nomographs were created using the “RMS” software to predict the disease progression rates after 12, 18, and 24 months. The calibration curve showed the performance of the nomograms with the observed rates at 12, 18, and 24 months.

The optimal cutoff value of the risk score was calculated using the R package “maxstat”. We set the minimum number of samples in each group to be greater than 25% and the maximum number of samples in each group to be less than 75%. Patients were further classified into high or low-risk groups based on this criterion with a cutoff of 0.42. The prognosis difference between the two groups was estimated using the “Survfit” function in the “survival” package and the log-rank test. Finally, we obtained the area under the curve (AUC) by receiver operating characteristic (ROC) analysis using the R package “pROC”. In particular, we gathered the patients’ follow-up durations and risk scores and performed the ROC analyses at 12, 18, and 24 months.

### Statistical and bioinformatic analysis

Before our data analysis, the protein matrix was log_2_-transformed. The coefficients of variation (CVs) of the proteins across the pooled samples and the Spearman correlation coefficients between pairs of pooled samples were then used to evaluate the data quality. Specifically, missing values were excluded from calculating the CVs and correlations. The R package “limma” was used for selecting the DEPs. Pathway and gene ontology (GO) enrichment were performed using Ingenuity Pathways Analysis (IPA 21.0) and Cytoscape (v3.8.1) with the ClueGO plugin (v2.5.7).

## Results

### Study design and global view of our proteomics analysis

To stratify the patients with long- or short-term progression from HSPC to CRPC, we collected and analyzed two sets of pre-ADT PCa samples (n=78): a discovery set (n=16) and a modeling set (n=62). The whole study was divided into four stages: 1) processing the FFPE- PCT-PulseDIA pipeline; 2) exploring the proteomics characteristics and biological differences between long- and short- term progression sample groups; 3) modeling for patient stratification; 4) progression and prognosis prediction (**Figure 1A**).

**Figure 1.**
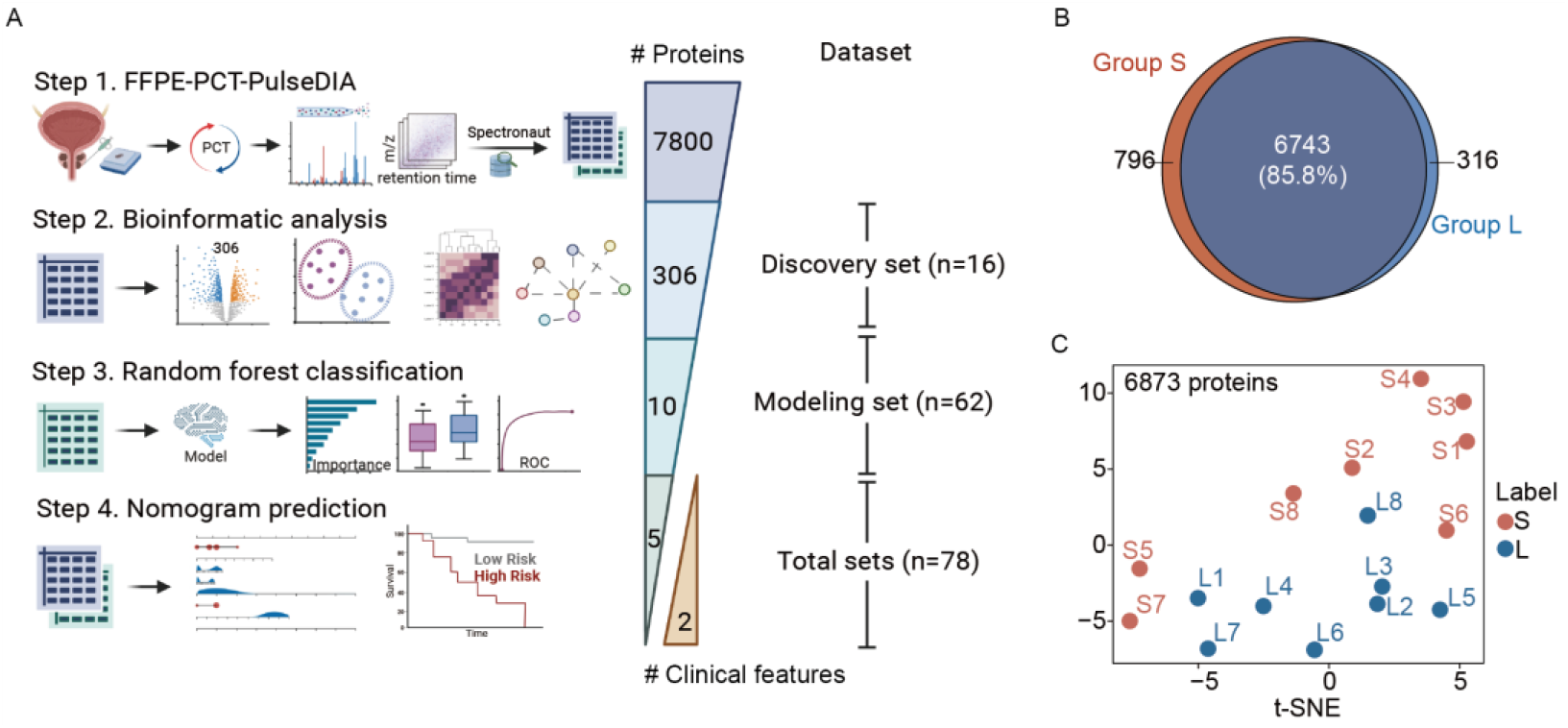
Study design and global proteomics view of the discovery set. (A) Workflow of the study. (B) Venn diagram of the protein identifications from the short- (S) and long-term (L) progression groups. (C) t-SNE plot showing the sample distribution of the S and L groups based on 6,873 proteins with less than 85% missing values.

In the first stage, we compared the proteomics expression differences between the two main groups in the discovery set: the long- (Group L, n=8) and short-term (Group S, n=8) progression groups. Specifically, Group L had shown a progression of at least twenty months, while Group S had a progression of no more than eight months. This time refers to the interval between the diagnosis of PCa and the diagnosis of CRPC. A total of 66,450 peptides, 7,961 protein groups, and 7,855 proteotypic proteins were identified in the 16 samples (**Figure 1B and sFigure 1A**). An average of 34,805 peptides and 5,318 proteins were detected. In particular, the median numbers of peptides and proteins were 31,658 and 5,118 for Group L and 38,375 and 5,615 for Group S. A total of 85.8% (6,743/7,855) proteins was identified across both groups (**sFigure 1B**). In contrast, 796 and 316 proteins were exclusively expressed in Group S and Group L, respectively (**Figure 1B**). The higher number of unique proteins identified in Group S may be explained by the more severe condition of the disease.

Next, we kept the proteins with less than 85% missing values that were detected in at least three samples, resulting in 6,873 proteins with relatively high confidence. The two sample groups were partially separated in the global view of the t-SNE plot of the 6,873 protein features (**Figure 1C**). This result suggests biological differences between the two groups.

### Proteomics characteristics and differences between long- and short-term progression patients

To explore the biological difference between the long- and short-term progression patients, we first identified the dysregulated proteins and visualized them using a volcano plot (**Figure 2A**). This comparison revealed 43 down-regulated and 263 up-regulated proteins in Group S with fold-changes > 2 and *p*-values < 0.01 (**Figure 2A**). The two groups could be clearly distinguished in the t-SNE plot based on these 306 DEPs (**Figure 2B**). The heatmap in **Figure 2C** shows the expressions of the 38 most significantly dysregulated proteins.

**Figure 2.**
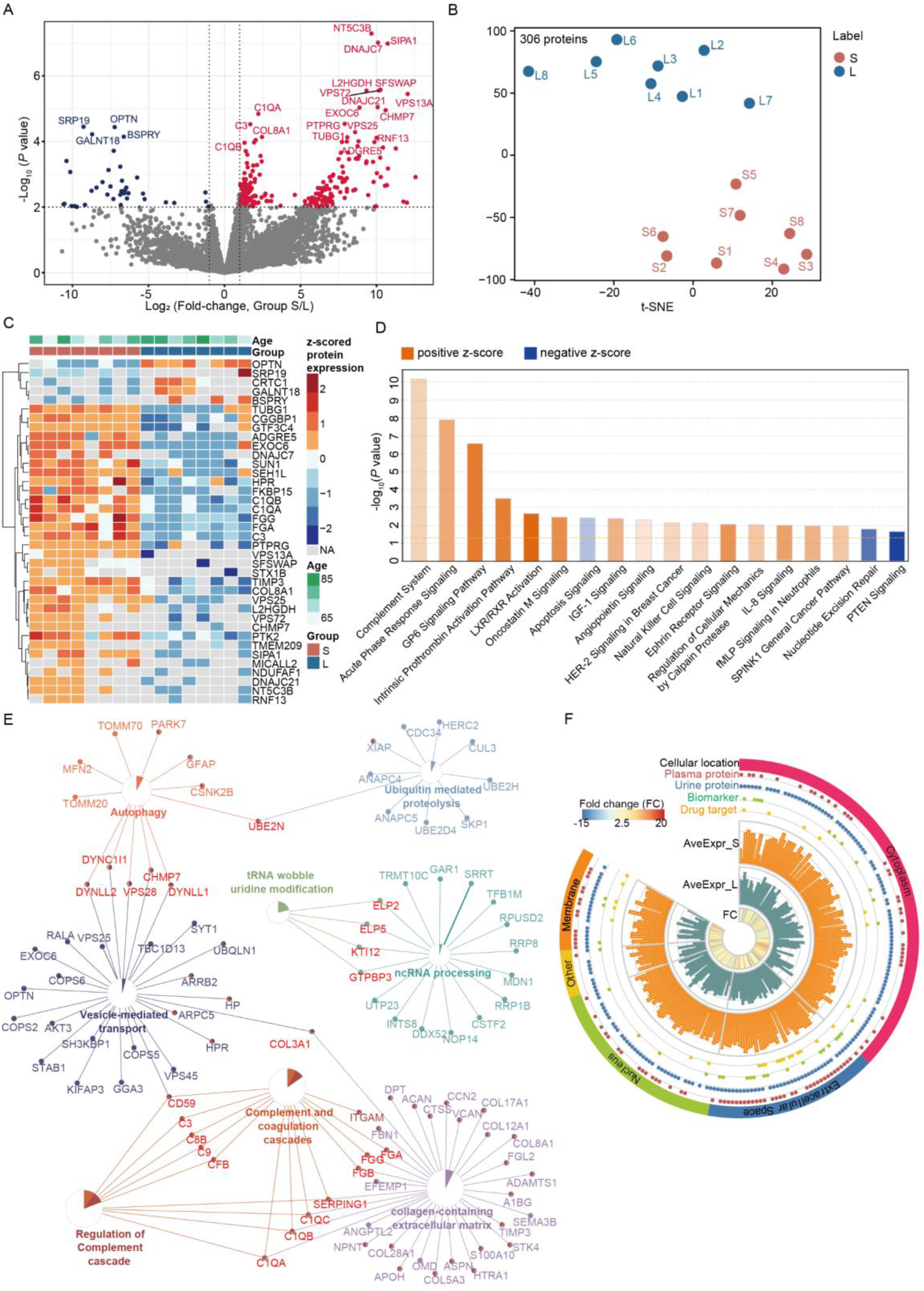
Comparative proteomics and bioinformatic analysis of the discovery set. (A) Volcano plot of the 306 differentially expressed proteins (DEPs) between Group S and Group L with *p*-value < 0.05 and fold-change > 2. (B) Using the 306 DEPs, the two groups were separated in the t-SNE plot. (C) Heatmap showing the expression of the 38 most significantly dysregulated proteins for each sample. (D) Pathway enrichment of the 306 DEPs. Positive and negative z-scores indicate the active and inhibited pathways, respectively. (E) The gene ontology enrichment networks of the 306 DEPs generated by the ClueGO plugin of Cytoscape. (F) Ciros plot showing the 306 DEPs annotated by cellular location, their being plasma proteins, urine proteins, biomarkers, and/or drug targets, their average expression in Group S (AveExpr_S) and Group L (AveExpr_L), and their fold change (FC).

Next, the 306 DEPs were enriched using pathway and GO analyses. The signaling pathways of the complement system, the acute phase response, and GP6 were the three most significantly activated in S group. Also, the pathways involved in apoptosis, nucleotide excision repair, and PTEN signaling were inhibited in Group S (**Figure 2D**). Our GO enrichment analysis showed that, except for the complement system, several biological processes are involved in an unfavorable prognosis of the disease: collagen-containing extracellular matrix, vesicle-mediated transport, autophagy, ubiquitin mediated proteolysis, ncRNA processing, and tRNA wobble uridine modification (**Figure 2E**). The above data revealed the most important signaling pathways, biological processes, and the corresponding proteins that are associated with a rapid disease progression. The dysregulated proteins we identified were detected in multiple biofluids, and they could potentially be used as diagnosis or prognosis biomarkers and therapeutic drug targets (**Figure 2F**).

### Identifying patients with rapid progression using a protein panel-based machine learning model

To stratify the two groups using statistical models, we collected a larger sample set: the modeling set (n=62). Using these samples and the same analytical methods used for the discovery set, we identified 7,423 proteins; the quality control analysis proved the data quality to be satisfactory (median CV = 0.0377; correlation coefficients > 0.85) (**sFigure 1C-1D**). Within the training set (n = 41), we identified the most significant proteins by setting the ANOVA *p*-values < 0.05 and then selected ten proteins with statistically significant differences. Among these, five proteins were significantly downregulated, and the other five were significantly upregulated in Group S (*p*-value < 0.05) (**Figure 3A**). We then built our RF model and ranked these ten protein features based on their Gini importance in our model (**Figure 3B**). The model performance on training set was shown in **sFigure 2**. Next, each individual in the testing set (n =21) was scored by the RF model based on the above ten features (**Figure 3C**). The resulting model correctly classified 18 of 21 patients (testing set) with an accuracy of 0.86 (**Figure 3D**); the model’s sensitivity, specificity, positive predictive value, and negative predictive value were 0.91, 0.80, 0.83, and 0.89, respectively. The AUC of our RF model was 0.914 (**Figure 3E**), showing the high performance of our classification model.

**Figure 3.**
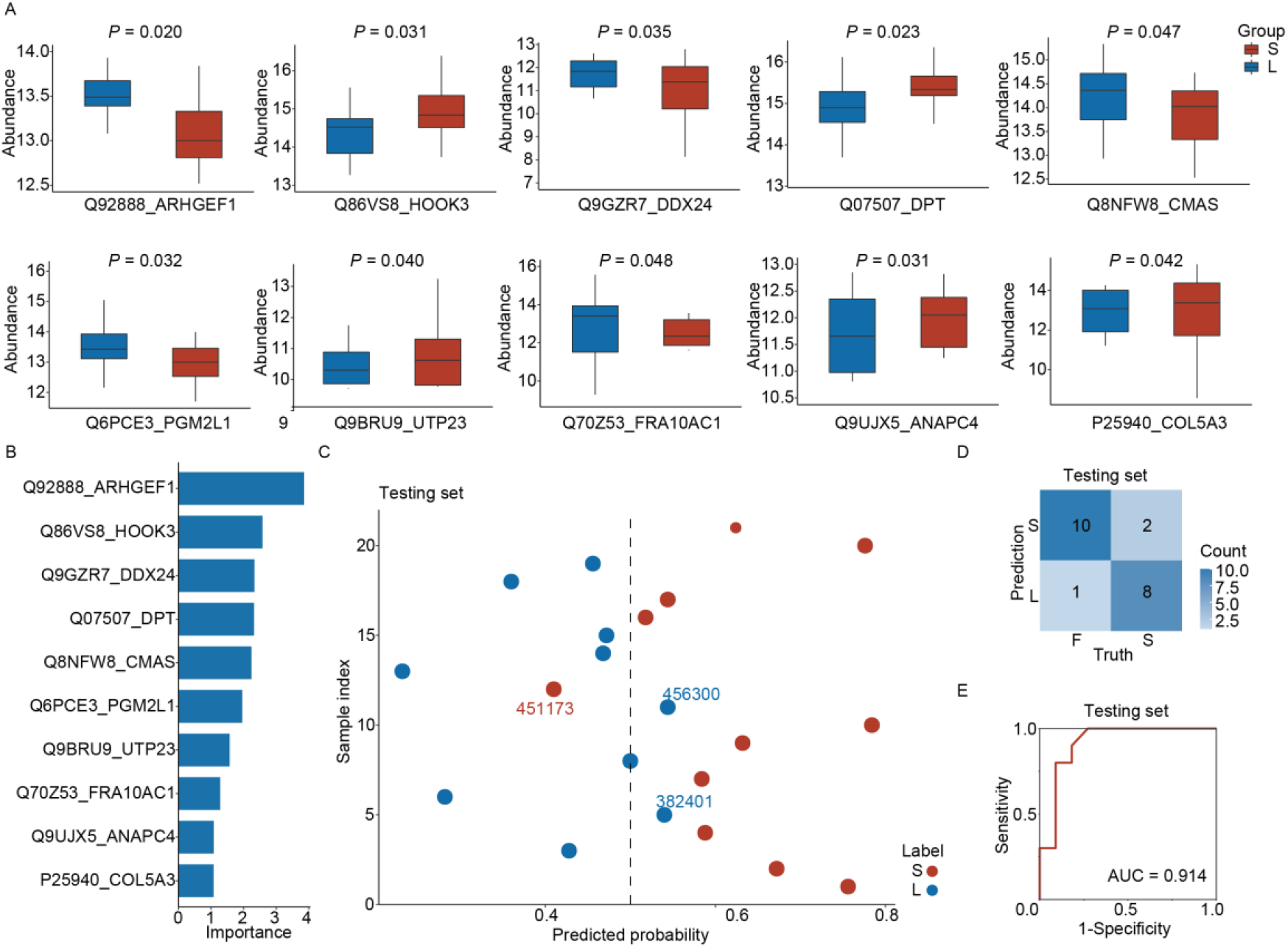
Stratification and prediction for patients with short- and long-term progression. (A) Boxplots showing the log_2_(protein abundance) in Groups S and L. The *p*- values were estimated using ANOVA. (B) The Gini importance of each protein feature ranked by our random forest model. (C) The predicted probabilities of belonging to Groups S or L for the patient from our testing set. Our random forest model generated these results. The three wrongly classified patients were marked with their patient identification numbers. (D) Confusion matrix of the independent testing set. (E) ROC plot showing the model performance with an independent testing set.

### Predicting the progression time and risk ratios through nomogram

We next predicted the progression time and the risk of progression to CRPC for each individual. To this aim, we screened the clinical characteristics and the above ten protein features using a multivariable Cox proportional hazards regression. Seven of the 16 features were significantly associated with an unfavorable prognosis. Specifically, two clinical features (GS and tPSA) and four protein features (DPT, ARGEF1, UTP23, and ANAPC4) were positively associated with the disease progression. In contrast, the most significantly modulated protein CMAS was negatively related to the progression and had a *p*-value = 0.0018 **(Figure 4A**). Hence, these seven features may help predict the probability of individual PCa cases progressing into CRPC.

**Figure 4.**
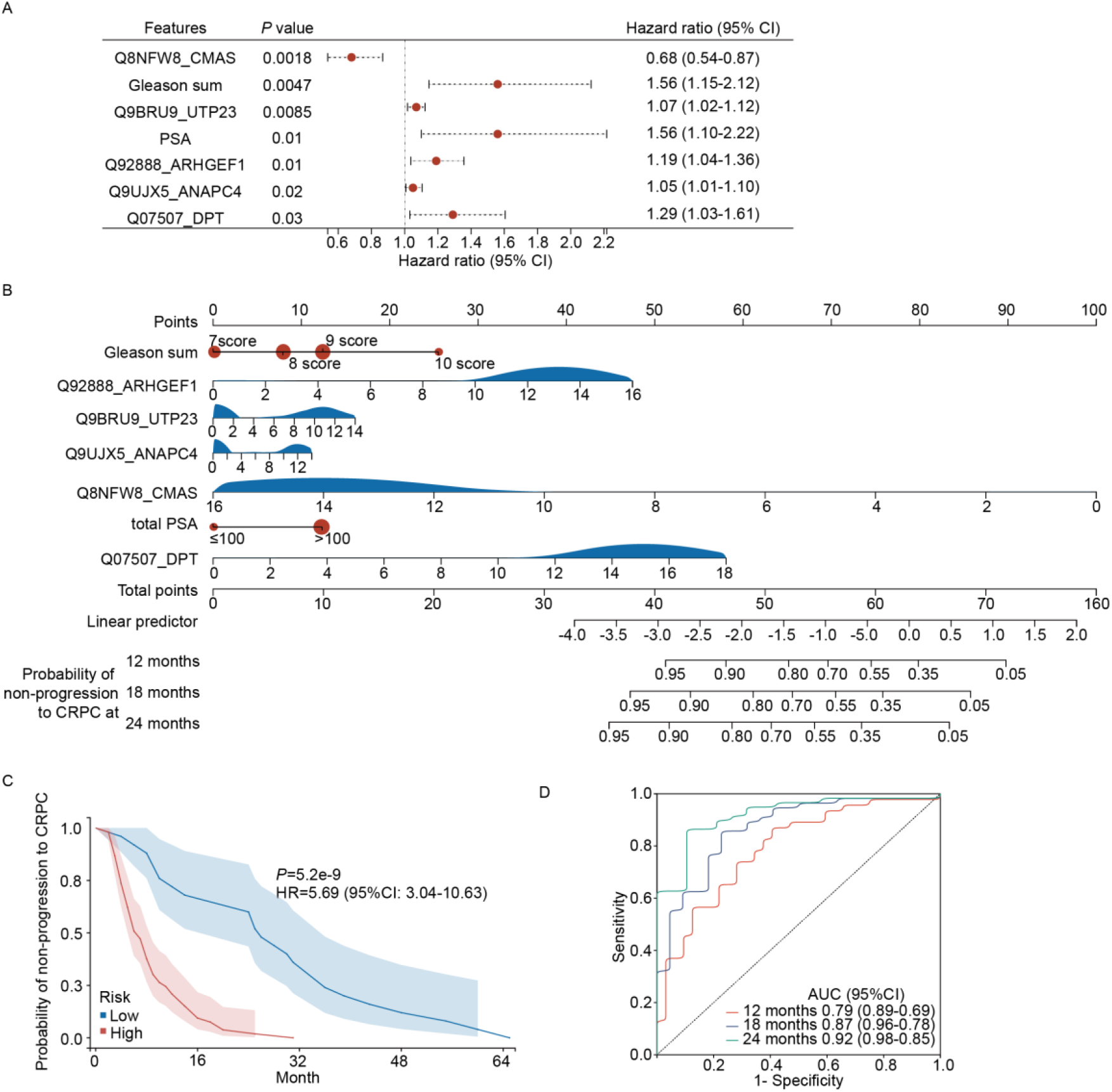
Predicting the probability of non-progression to castration-resistant prostate cancer (CRPC) at different times. (A) Multivariable Cox analysis of the proteins and clinical features we selected for distinguishing progression to CRPC (*p*-value < 0.05). (B) Nomogram for the prognostic prediction of developing advanced hormone-sensitive prostate cancer. (C) Survival curves indicating the probability of non-progression to CRPC. The low- and high-risk are grouped using an optimal nomogram risk score cutoff of 0.42. (D) ROC curves for the 12-, 18-, and 24-month advanced prostate cancer progress-related nomogram.

Next, we constructed a nomogram by integrating these seven features and predicted the disease progression (**Figure 4B**). Each individual was scored using our nomogram and the seven features, with a risk score cutoff of 0.42. Specifically, based on this cutoff, patients were split into groups of high- (n = 25, ratio of 32%) and low-risk scores (n=53, ratio of 68%). The model’s overall concordance index (C-index) was 0.72 (95% CI 0.66-0.78), and our calibration plot showed the agreement between our estimations and the observations 18 and 24 months after confirmed HSPC. The progression curve showed a statistically significant difference between the two groups (*p-*value = 5.2 × 10^−9^, HR = 5.69) (**Figure 4C**). The AUC values of the 18-month (AUC=0.87) and 24-month (AUC=0.92) nomogram progressions were larger than those of the 12-month (AUC=0.79) ones (**Figure 4D**). Therefore, the most accurate nomograms for predicting the development of PCa were the 18-month and 24-month nomograms.

## Discussion

We investigated pre-ADT PCa patients by exploring the differences between the long- and short-term progression cases. We collected clinical data and measured the proteomes of PCa biopsies. We newly identified proteins that may be crucial in a faster progression of HSPC into CRPC. Finally, we generated a model for predicting the advancement of HSPC and stratifying patients into high- and low-risk groups.

The clinical staging of newly diagnosed PCa patients in China differs from western developed countries. For instance, among the newly diagnosed PCa patients in the US, most cases (∼76%) are clinically localized, while only ∼13% and ∼6% involve metastases in local lymph nodes or distant sites, respectively^16^. However, the data in China are quite different. A multi-center Chinese study showed that only 1/3 of the newly diagnosed PCa patients are clinically localized. Also, most patients in China are in the middle or advanced stage at the diagnosis, resulting in a worse overall prognosis than in Western countries^17^. For this reason, our study focused on exploring advanced PCa cases from China. In our study, we enrolled 78 advanced PCa patients, but their median time for progressing to CRPC was only nine months: much shorter than previously published^18^. This fact may be explained by most of our enrolled patients already having developed metastases (N = 72, 97.3%).

Recent studies have shown that the time of HSPC progression to CRPC is highly variable in patients treated with standard ADT^19^. Multiple phase-III trials proved the importance of identifying HSPC patients at risk of rapid disease progression, allowing the early implementation of appropriate therapeutic strategies to improve the prognosis^20^. There are two well-known studies on the risk classification of metastatic HSPC. The first was the CHAARTED trial, where patients with visceral and/or at least four bone metastases were classified as high-volume to distinguish them from the remaining low-volume ones^21^, concluding that the high-volume group benefits from ADT+ docetaxel treatment, whereas the low-volume group should be served by ADT alone. The second study was the LATITUDE trial, where patients with at least two high-risk characteristics (at least three bone metastases, visceral metastases, and ISUP grade four) were categorized as high-risk, which were found to have an increased survival following an abiraterone acetate plus prednisone therapy^7^. However, it is obvious that there were many patients defined as low-risk or low-volume, progressed to CRPC rapidly in the above researches. And in our study, most patients would have been classified as low-volume and low-risk according to the CHAARTED and LATITUDE criteria, respectively. However, our patients had a rapid disease progression (median time: nine months). Collectively, stratifying patients based on clinical imaging or M stage alone is inappropriate.

In this study, a novel nomogram was established for predicting the probability of specific progression times to CRPC, by integrating five proteins (ARHGEF1, UTP23, ANAPC4, DPT, and CMAS) and two clinical features (tPSA and GS), based on their expression levels and regression coefficients, as demonstrated by the multivariate Cox regression analysis. As a comprehensive scoring system, of which prediction ability was comfirmed by the overall C-index and the AUC values, our results suggest a better discrimination capability than previously published transcriptomic signatures consisting of two to 22 genes^22-25^.

According to the risk score, patients were then divided into high-risk and low-risk groups. The Kaplan–Meier survival curve showed that patients with high-risk scores had a significantly poorer recurrence-free survival than those with low-risk scores, suggesting that patients with high-risk scores were more prone to progression. According to current clinical trails mentioned above, high-risk patients may respond better to ADT combined with chemotherapy or novel endocrine therapy medicines. Larger-scale clinical trials are necessary to assess the most suitable treatments for each identified category.

In terms of the five proteins in the model, ARHGEF1 and DPT have been proved to promote the occurrence and progression of other cancers^26-28^, which were consistent with our results. ANAPC4 and CMAS have not been reported in cancer. It is noteworthy that down-regulation of UTP23 promotes paclitaxel resistance and predicts poorer prognosis in ovarian cancer^29^. However, in the present study, high expression of UTP23 promoted the progressions from HSPC to CRPC. Subsequently, we looked at the effect of UTP23 on the prognosis of pan-cancer through the TCGA database. The effect of UTP23 on the prognosis of different cancer types was different, including significantly promoting cancer in 7 cancer types and suppressing cancer in 2 cancer types. In TCGA database, the trend of UTP23 promoting prostate cancer is consistent with our study. The role of UTP23 in different cancer types deserves further study.

Although several proteomics studies of PCa have been published, they mainly focused on characterizing protein alterations and their biological changes by comparing benign/normal with PCa cases^30^, exploring metastatic PCa^31^, describing the heterogeneity of PCa^32^, and investigating the disease progression^8^. Ours is the first study to employ proteomics to investigate the differences between long- and short-term progressions from HSPC to CRPC and to discover a panel biomarker for identifying patients with a rapid progression. The ∼8000 proteins we identified provide a high-quality resource for explorative analyses. Furthermore, the 306 DEPs we found associated with different progressions to CRPC were enriched in the complement system and other inflammatory response-related pathways and functions **(Figure 2D and 2E)**, in agreement with previous findings using prostatic fluids^33^. In GO enrichment analysis, collen-containing extracellular matrix was closely related to DPT proteins. The changes of collen-containing extracellular matrix caused by high expression of DPT will become a potential research direction for HSPC progression to CRPC.

Our study was limited by the small sample size, which was collected from a single center. Also, all our patients were treated with standard ADT. However, in clinical practice, ADT, combined with bicalutamide and flutamide, is gradually being replaced by the combination of chemotherapy and second-generation hormonotherapy. Therefore, further validations need to be performed on a larger and multi-center study. In particular, individuals with different treatment approaches should be compared to determine the benefits of various treatment strategies in connection with patient stratification.

## Conclusion

We identified proteins and clinical parameters that are significantly associated with a fast progression of HSPC to CRPC. Using this information, we developed efficient models for classifying PCa patients and predicting HSPC development. Our data and tools can potentially guide the clinical management of patients with advanced HSPC.

## Data Availability

The mass spectrometry proteomics data have been deposited to the iProX with the dataset identifier IPX0005031000 (the data will be publicly released upon publication).

## Conflict of interest

T.G. is a shareholder of Westlake Omics, Inc. L.H. and W.G. are employees of Westlake Omics Inc, C.P., Y.H., H.W., Y.Y., L.L., M.L., B.Y., Y.S., T.G., and Z.L. have applied for a patent on this project. The remaining authors declare no competing interest.

## Acknowledgments

We thank Yingrui Wang, Xuan Ding and Jun A from Westlake University for the modification suggestions which help with improving the manuscript. This work is supported by grants from the National Key R&D Program of China (No. 2021YFA1301601, 2021YFA1301602, 2020YFE0202200) to T.G.; “1+X” program for Clinical Competency enhancement–Clinical Research Incubation Project and the Second Hospital of Dalian Medical University (2022LCYJZD02) to B.Y.; United Fund of the Second Hospital of Dalian Medical University and Dalian Institute of Chemical Physics and Chinese Academy of Sciences (UF-ZD-202014) to Y.Y.

## Author contributions

Z.L., B.Y., Y.H. and Y.S. designed the project. Y.H. and Y.Y. collected the FFPE biopsies. Y.S. and L.L. performed the experiments for proteomics preparation. L.H., M.L. and W.G. conducted proteomics data analysis. H.W. constructed the random forest model. C.P. performed clinical data analysis and nomogram analysis. Y.H., C.P., H.W., and Y.S. wrote the manuscript with inputs from all co-authors. Y.S. T.G. and Z.L. supervised the project.

## Supplementary Materials

**sTable 1.**
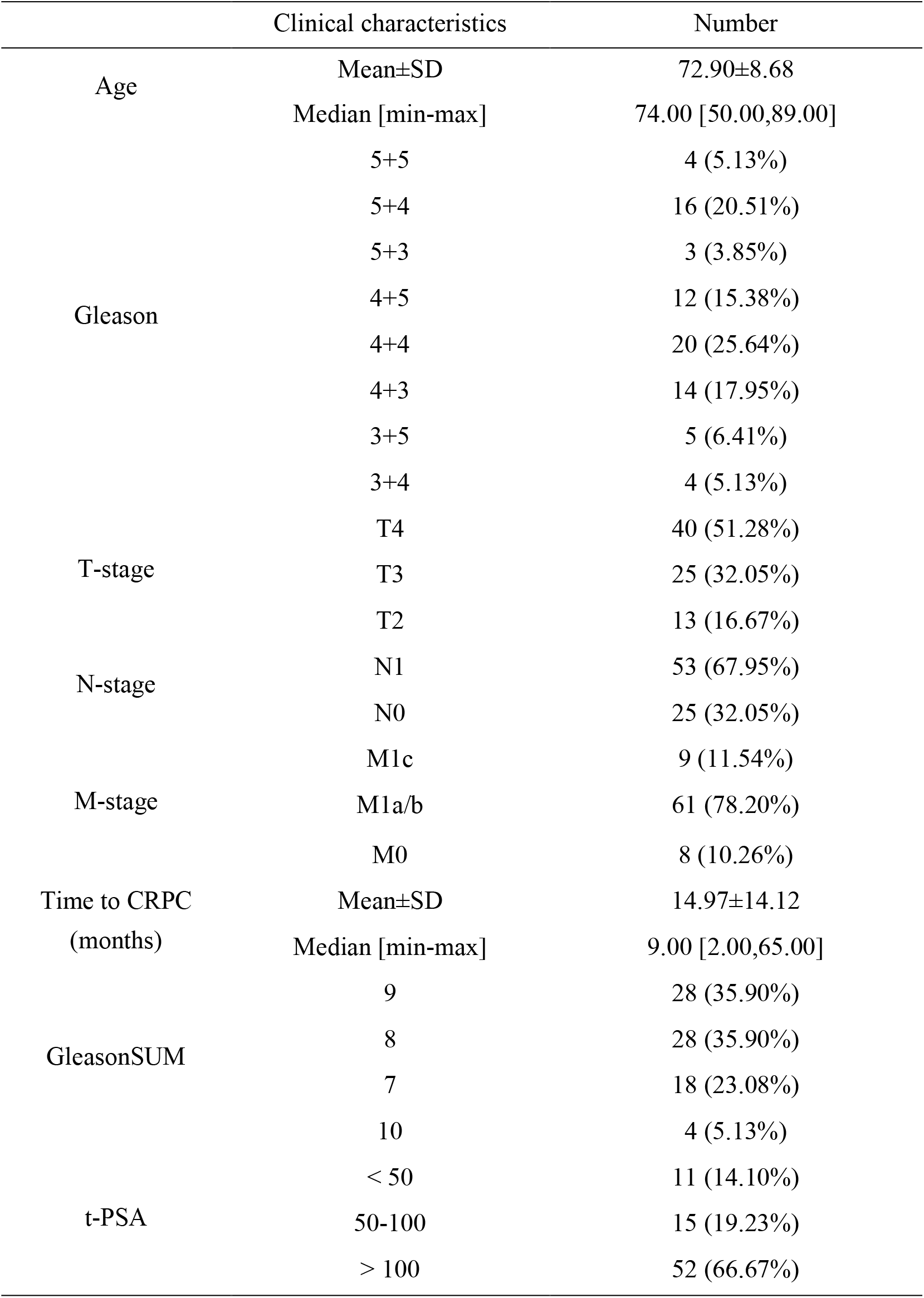
Representativeness of Study Participants.

**sFigure 1.**
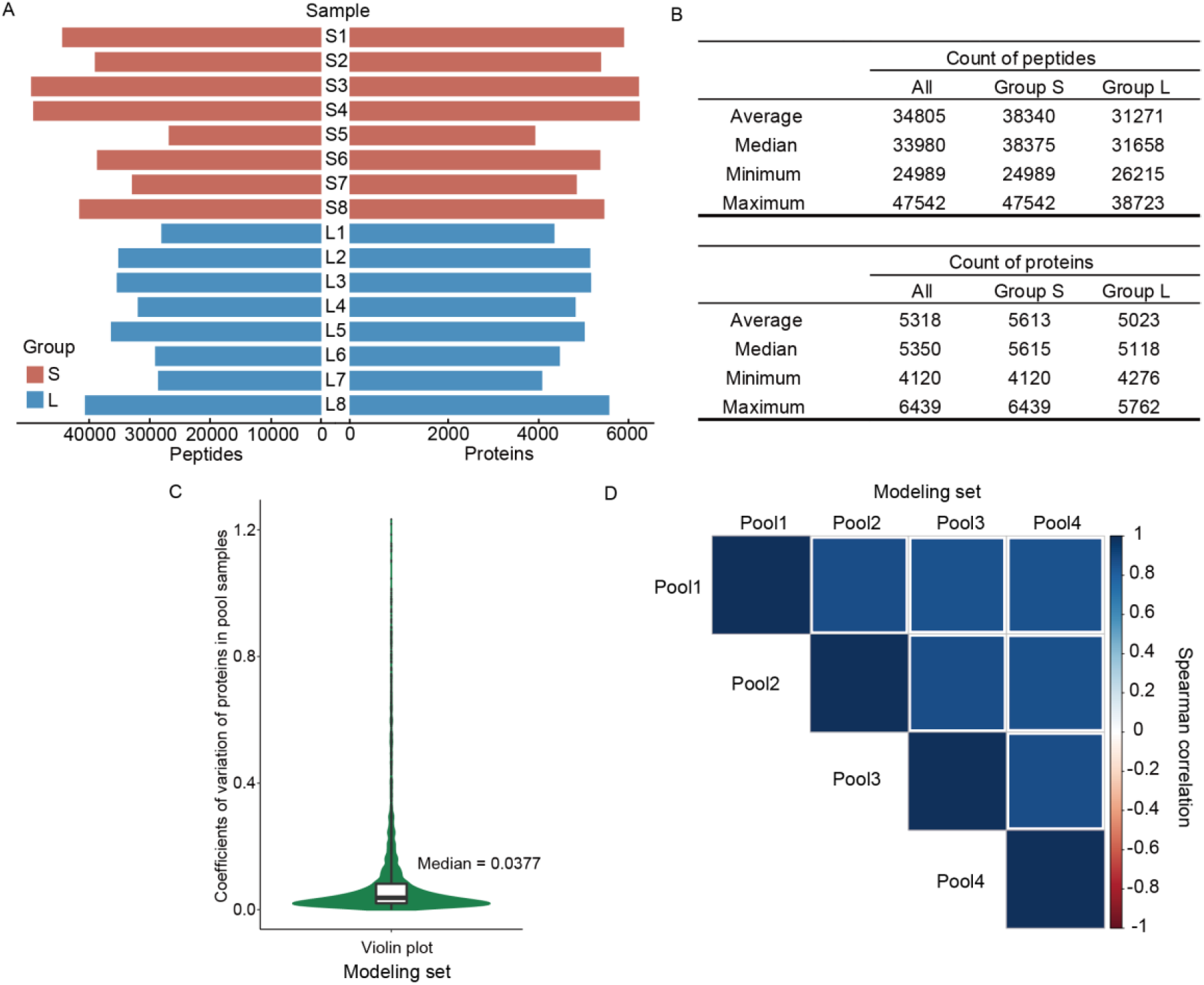
Data quality control analysis. (A) Numbers of the identified proteins and peptides from Group F and Group S in the discovery set. (B) Statistical analysis for the peptide and protein identifications in the discovery set. (C) Coefficients of variation of the proteins across the pooled samples in the modeling set. (B) Pearson correlations between pairs of pooled samples in the modeling set.

**sFigure 2.**
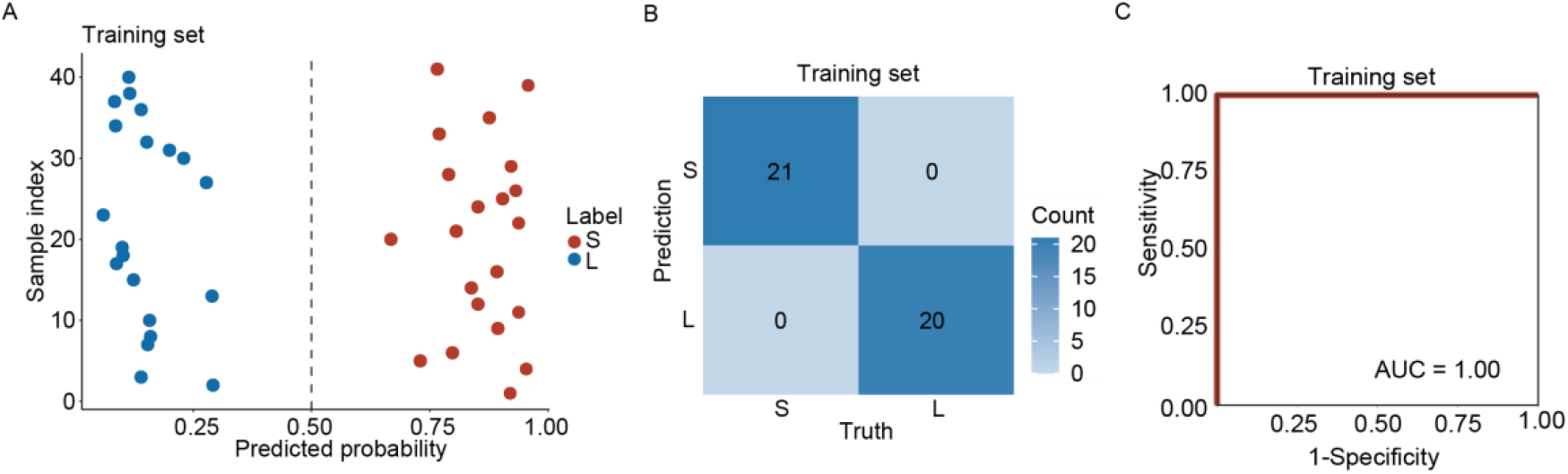
Model performance with the training set. (A) The predicted probabilities of belonging to Groups S or L for the patient in our training set. Our random forest model generates these results. (B) The confusion matrix of the training set. (C) The receiver operating characteristic curve of the training set.

**sFigure 3.**
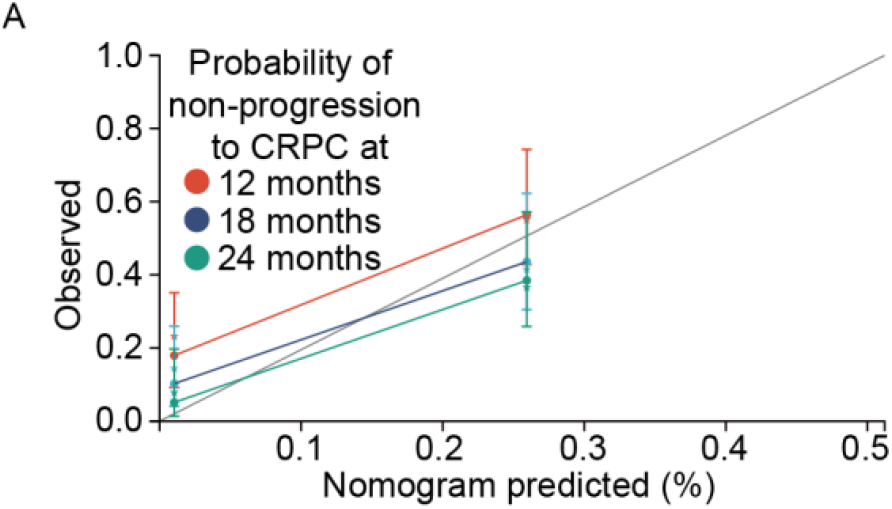
The calibration curves of our nomogram. (A) Calibration curves for the 12-, 18-, and 24-month progressions to advanced prostate cancer.

## Notes

### Funding Statement

This study was funded by grants from the National Key R&D Program of China (No. 2021YFA1301601, 2021YFA1301602, 2020YFE0202200) to T.G.; 1+X program for Clinical Competency enhancement Clinical Research Incubation Project and the Second Hospital of Dalian Medical University (2022LCYJZD02) to B.Y.; United Fund of the Second Hospital of Dalian Medical University and Dalian Institute of Chemical Physics and Chinese Academy of Sciences (UF ZD 202014) to Y.Y.

### Author Declarations

Ethics committee of the Second Hospital of Dalian Medical University gave ethical approval for this work.

## References

1 Sung, H. et al. Global Cancer Statistics 2020: GLOBOCAN Estimates of Incidence and Mortality Worldwide for 36 Cancers in 185 Countries. CA Cancer J Clin 71, 209–249, doi:10.3322/caac.21660 (2021).

2 Chen, W. et al. Cancer statistics in China, 2015. CA Cancer J Clin 66, 115–132, doi:10.3322/caac.21338 (2016).

3 Shaw, P. J. & Feske, S. Physiological and pathophysiological functions of SOCE in the immune system. Front Biosci (Elite Ed) 4, 2253–2268, doi:10.2741/e540 (2012).

4 Bjartell, A. Circulating tumour cells as surrogate biomarkers in castration-resistant prostate cancer trials. Eur Urol 60, 905–907, doi:10.1016/j.eururo.2011.08.024 (2011).

5 Harris, W. P., Mostaghel, E. A., Nelson, P. S. & Montgomery, B. Androgen deprivation therapy: progress in understanding mechanisms of resistance and optimizing androgen depletion. Nat Clin Pract Urol 6, 76–85, doi:10.1038/ncpuro1296 (2009).

6 Kyriakopoulos, C. E. et al. Chemohormonal Therapy in Metastatic Hormone-Sensitive Prostate Cancer: Long-Term Survival Analysis of the Randomized Phase III E3805 CHAARTED Trial. J Clin Oncol 36, 1080–1087, doi:10.1200/JCO.2017.75.3657 (2018).

7 Fizazi, K. et al. Abiraterone acetate plus prednisone in patients with newly diagnosed high-risk metastatic castration-sensitive prostate cancer (LATITUDE): final overall survival analysis of a randomised, double-blind, phase 3 trial. Lancet Oncol 20, 686–700, doi:10.1016/S1470-2045(19)30082-8 (2019).

8 Latonen, L. et al. Integrative proteomics in prostate cancer uncovers robustness against genomic and transcriptomic aberrations during disease progression. Nat Commun 9, 1176, doi:10.1038/s41467-018-03573-6 (2018).

9 Guo, T. et al. Rapid mass spectrometric conversion of tissue biopsy samples into permanent quantitative digital proteome maps. Nat Med 21, 407–413, doi:10.1038/nm.3807 (2015).

10 Zhu, Y. et al. High-throughput proteomic analysis of FFPE tissue samples facilitates tumor stratification. Mol Oncol 13, 2305–2328, doi:10.1002/1878-0261.12570 (2019).

11 Mottet, N. et al. EAU-ESTRO-SIOG Guidelines on Prostate Cancer. Part 1: Screening, Diagnosis, and Local Treatment with Curative Intent. Eur Urol 71, 618–629, doi:10.1016/j.eururo.2016.08.003 (2017).

12 Cornford, P. et al. EAU-ESTRO-SIOG Guidelines on Prostate Cancer. Part II: Treatment of Relapsing, Metastatic, and Castration-Resistant Prostate Cancer. Eur Urol 71, 630–642, doi:10.1016/j.eururo.2016.08.002 (2017).

13 Cai, X. et al. High-throughput proteomic sample preparation using pressure cycling technology. Nat Protoc, doi:10.1038/s41596-022-00727-1 (2022).

14 Cai, X. et al. PulseDIA: Data-Independent Acquisition Mass Spectrometry Using Multi-Injection Pulsed Gas-Phase Fractionation. J Proteome Res 20, 279–288, doi:10.1021/acs.jproteome.0c00381 (2021).

15 Sun, Y. et al. Stratification of follicular thyroid tumours using data-independent acquisition proteomics and a comprehensive thyroid tissue spectral library. Mol Oncol 16, 1611–1624, doi:10.1002/1878-0261.13198 (2022).

16 Siegel, R. L., Miller, K. D., Fuchs, H. E. & Jemal, A. Cancer Statistics, 2021. CA Cancer J Clin 71, 7–33, doi:10.3322/caac.21654 (2021).

17 Ma, C. G. et al. [Epidemiology of prostate cancer from three centers and analysis of the first-line hormonal therapy for the advanced disease]. Zhonghua Wai Ke Za Zhi 46, 921–925 (2008).

18 Sumanasuriya, S. & De Bono, J. Treatment of Advanced Prostate Cancer-A Review of Current Therapies and Future Promise. Cold Spring Harb Perspect Med 8, doi:10.1101/cshperspect.a030635 (2018).

19 Napoli, G. et al. A Systematic Review and a Meta-analysis of Randomized Controlled Trials’ Control Groups in Metastatic Hormone-Sensitive Prostate Cancer (mHSPC). Curr Oncol Rep, doi:10.1007/s11912-022-01323-y (2022).

20 Barata, P. C. & Sartor, A. O. Metastatic castration-sensitive prostate cancer: Abiraterone, docetaxel, or. Cancer 125, 1777–1788, doi:10.1002/cncr.32039 (2019).

21 (!!! INVALID CITATION !!! 7).

22 Li, S. et al. Establishment of a Novel Combined Nomogram for Predicting the Risk of Progression Related to Castration Resistance in Patients With Prostate Cancer. Front Genet 13, 823716, doi:10.3389/fgene.2022.823716 (2022).

23 A J. Zhang, B., Zhang, Z., Hu, H. & Dong, J. T. Novel Gene Signatures Predictive of Patient Recurrence-Free Survival and Castration Resistance in Prostate Cancer. Cancers (Basel) 13, doi:10.3390/cancers13040917 (2021).

24 Wang, Y. & Yang, Z. A Gleason score-related outcome model for human prostate cancer: a comprehensive study based on weighted gene co-expression network analysis. Cancer Cell Int 20, 159, doi:10.1186/s12935-020-01230-x (2020).

25 Hu, D. et al. Development of an autophagy-related gene expression signature for prognosis prediction in prostate cancer patients. J Transl Med 18, 160, doi:10.1186/s12967-020-02323-x (2020).

26 Hasan, M. K. et al. Wnt5a induces ROR1 to recruit cortactin to promote breast-cancer migration and metastasis. NPJ Breast Cancer 5, 35, doi:10.1038/s41523-019-0131-9 (2019).

27 Guo, Y. et al. Dermatopontin inhibits papillary thyroid cancer cell proliferation through MYC repression. Mol Cell Endocrinol 480, 122–132, doi:10.1016/j.mce.2018.10.021 (2019).

28 Xi, L. C. et al. Effects of Dermatopontin gene silencing on apoptosis and proliferation of osteosarcoma MG63 cells. Mol Med Rep 17, 422–427, doi:10.3892/mmr.2017.7866 (2018).

29 Fu, Z. et al. Down-regulation of UTP23 promotes paclitaxel resistance and predicts poorer prognosis in ovarian cancer. Pathol Res Pract 215, 152625, doi:10.1016/j.prp.2019.152625 (2019).

30 Iglesias-Gato, D. et al. The Proteome of Primary Prostate Cancer. Eur Urol 69, 942–952, doi:10.1016/j.eururo.2015.10.053 (2016).

31 Iglesias-Gato, D. et al. The Proteome of Prostate Cancer Bone Metastasis Reveals Heterogeneity with Prognostic Implications. Clin Cancer Res 24, 5433–5444, doi:10.1158/1078-0432.CCR-18-1229 (2018).

32 Guo, T. et al. Multi-region proteome analysis quantifies spatial heterogeneity of prostate tissue biomarkers. Life Sci Alliance 1, doi:10.26508/lsa.201800042 (2018).

33 Grayhack, J. T., Wendel, E. F., Oliver, L. & Lee, C. Analysis of specific proteins in prostatic fluid for detecting prostatic malignancy. J Urol 121, 295–299, doi:10.1016/s0022-5347(17)56760-9 (1979).

